# Estimating emergence rates of epidemiologically relevant traits in bacteria with EMERGENe

**DOI:** 10.64898/2026.09.10.26362639

**Authors:** Gherard Batisti Biffignandi, Kuangyi Charles Wei, Joel Hellewell, Claire Jenkins, Jukka Corander, John A Lees, Kate S Baker

## Abstract

Genomic surveillance has transformed our ability to identify and track bacterial lineages and epidemiologically relevant traits. However, surveillance approaches are mainly based on prevalence-based measures, which can obscure the dynamics of traits undergoing rapid expansion within genetically and temporally heterogeneous populations. This limitation is particularly relevant for antimicrobial resistance (AMR), where the emergence and subsequent expansion of newly acquired transmissible traits can generate substantial changes in population-level prevalence. Here, we introduce EMERGENe (https://github.com/gbatbiff/EMERGENe), a phylogenetic framework that combines ancestral-state reconstruction and analysis of phyletic patterns to quantify the emergence dynamics of gained and transmissible traits. Using time-scaled phylogenies and binary presence data, EMERGENe identifies independent phyletic events in which a trait is acquired and subsequently inherited by its descendants, and estimates interpretable Entry and Emergence rates that capture the introduction and expansion of trait-specific populations. We evaluated the performance of EMERGENe using phylogenetic simulations across evolutionary trajectories with different population growth dynamics, and applied our framework to a national genomic surveillance dataset comprising 3,745 *Shigella sonnei* isolates. Across simulated evolutionary scenarios, EMERGENe was superior to prevalence for discriminating traits undergoing rapid population growth from those with slower or no expansion. Applied to *S. sonnei*, our method detected previously known epidemiological acquisition of resistance to azithromycin, ciprofloxacin and third-generation cephalosporins, while providing information on their underlying emergence dynamics. Temporal analyses also revealed the progressive expansion of ceftriaxone resistance, overlapping with the increasing replacement of previously highly disseminated azithromycin resistance. EMERGENe also detected emerging and overlooked traits, including a recently described epidemiologically relevant phage–plasmid and the *qnrS1* gene. EMERGENe provides a complementary approach to genomic surveillance by shifting the focus from static trait prevalence towards the evolutionary processes underlying trait emergence and expansion. Thus, EMERGENe provides a robust quantification method for comparison of trait emergence, and by identifying early signals of rapidly emerging traits, also has the potential to improve longitudinal surveillance, facilitating earlier intervention in the onward transmission of AMR in bacterial populations.

## Introduction

Recent implementations of bacterial whole genome sequencing in routine public health surveillance have significantly improved our ability to track the spread of genetic traits of epidemiological interest among bacterial species. However, the epidemiology and dynamics of transmissible genes conferring phenotypic traits of interest e.g. Antimicrobial Resistance (AMR), remains poorly understood (1) and translates to several surveillance limitations. This is partly attributable to a lack of robust quantitation methods that reflect the natural population dynamics including the transmissible nature of many of the traits of interest, the non-independence of surveillance samples, and the fluctuating nature of the bacterial populations in which they transmit. Specifically, current AMR surveillance approaches mostly rely on reporting ‘prevalence’ (actually proportional presence) data like percentage of isolates that are resistant to a particular antimicrobial within a given sampling frame, such as a calendar year. This approach masks critical information that ignores the complex underlying multifactorial evolutionary, ecological, and epidemiological drivers of trait emergence (2), ignoring a trait that is emerging rapidly in a population due to a clonal effect or multiple introductions (Figure 1). It has long been understood in the field of epidemiology that prevalence is fundamentally inappropriate for highly dynamic denominator populations and trait emergence in bacterial populations should be measured with something closer to formal incidence measures, such as cases/person-years (3).

**Figure 1.**
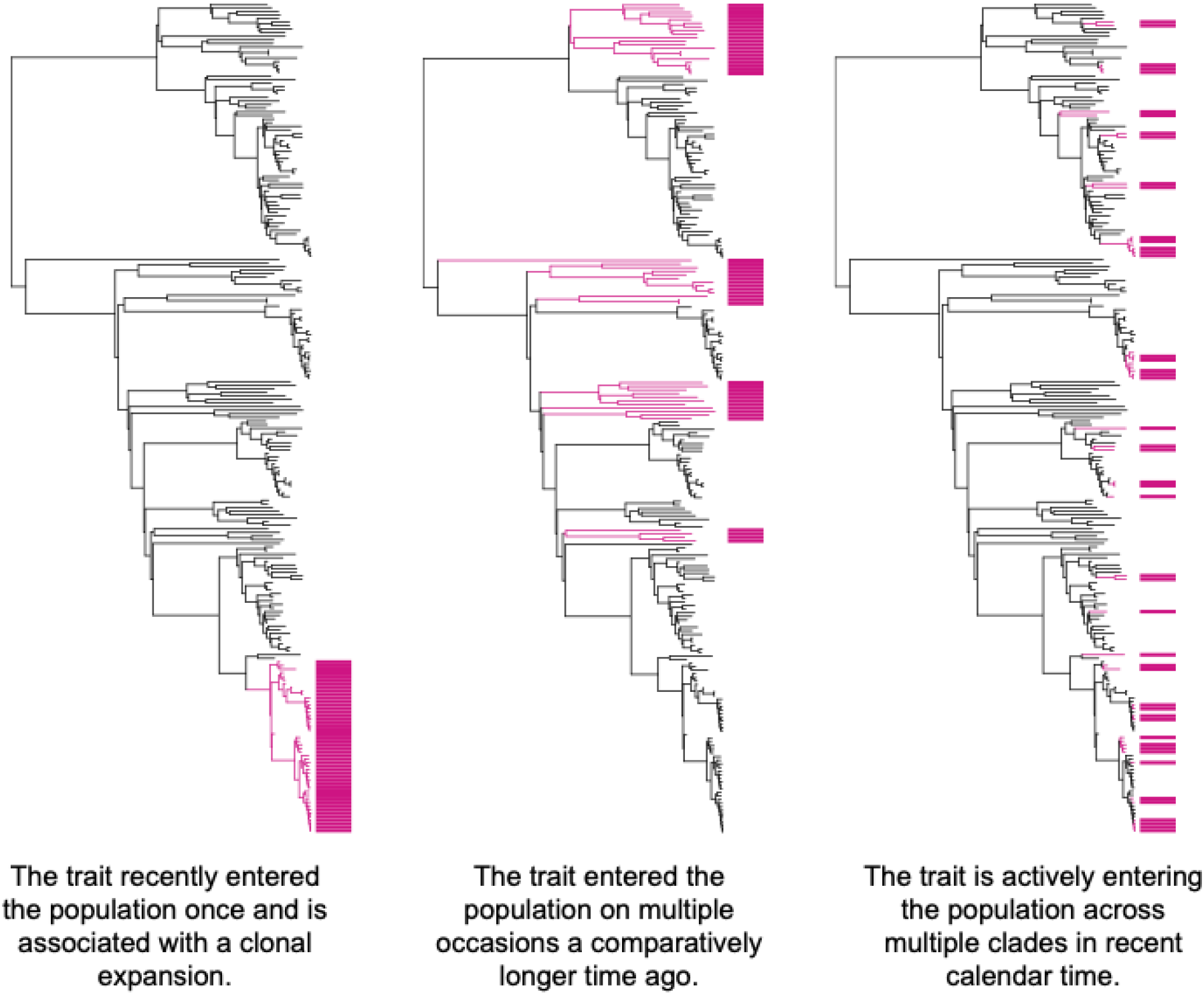
Variable ecological trajectories of an acquirable bacterial trait lost by proportional presence. The three time-tree schematics show example phylogenetic distributions of a trait (pink branches and metadata strip) across a tree comprising 231 isolates. In each case 21% (n=48) of isolates bear the trait, representing their uniform proportional presence value. The phylogenetic inferences written underneath each schematic describe the ecological trajectories which cannot be captured using proportional presence data.

The acquisition of genetic traits in bacteria is often driven by horizontal gene transfer (HGT) through mobile genetic elements (MGEs), such as plasmids (4,5). HGT events can often be observed in genomic data by their appearance in multiple clades within and across species (i.e. their being polyphyletic). The distribution of these features among bacteria can depend on several factors including selective pressures, host adaptation, and the fitness cost of their maintenance (6,7). Indeed, transmissible traits can be either beneficial or detrimental to their host, depending on the trade-off between their fitness cost and dynamic pressures exerted by the environment (8). As a result, transmissible traits subjected to selective events can result in subsequent spread and novel expansion of acquired traits within and across populations (9,10). New methods are needed to better identify signals of the early spread of traits in pathogens that could enable early intervention in, for example, combatting AMR.

Hallmarks of expansion growth are imprinted within phylogenetic shape (11), and can be detected through analyses such as early bursts of diversification, rapid branch splitting (12) and multiple mergers (9,13). It is also critical to analyse whether traits occur across multiple clades, as this polyphyly may indicate convergent evolution or multiple acquisitions of horizontally acquired traits being selected for in the bacterial population (14). For example, identical AMR plasmids have been observed to emerge polyphyletically and in distinct bacterial populations before fixation, and some studies suggest that measurable fitness costs may correlate with the epidemiological behaviour of plasmids (15–17). However, the absence of methods that allow for aggregated, repeatable quantification of polyphyletic traits prevent us from robustly linking and comparing phenotypes (e.g. plasmid fitness, AMR measures) with epidemiological emergence patterns.

Several phylodynamic models exist which were initially developed for viruses and more recently also applied to bacteria to infer population dynamics, e.g. estimating growth rates over time starting from genomic data (18–21). Several of these models are based on birth-death or coalescent theory (9,19,22–24), albeit many of them focused on the phylogenetic patterns (12), rather than incorporating additional information about discrete traits (e.g. gene/phenotype presence, ecological/geographical compartments) (25). Despite the potential importance of applying such models to infer the behaviour of biological traits, the application of phylogeography-based methods is less common in the context of bacteria, due to the complexity of their underlying population dynamics, variation in evolutionary rates across sites in the genome (incorporating the effect of mixed ecology), and often rapidly-fluctuating accessory genome content (24,26). This limitation can lead to the undetected spread of epidemiologically relevant traits, underlining a methodological gap critical for the early identification of AMR and emergence of other traits in genomic surveillance datasets.

Motivated by this capability gap for AMR surveillance, we developed EMERGENe, a novel phylogenetic method based on ancestral-state reconstruction and analyses of phyletic patterns to infer trait-specific population dynamics in bacteria. EMERGENe facilitates robust, quantitative comparison of trait behaviour and identification of early signals of associated clonal expansion. Using a timed tree and trait presence and absence data, our method returns epidemiologically relevant measures of: an Entry rate (how long the trait takes to fix in a clade), an Emergence rate (akin to incidence), and a count of Phyletic events of the trait(s) across the population. Here, we describe our method and demonstrate its utility and efficacy on both simulated and real data, including public health genomic surveillance datasets of the AMR-priority pathogen *Shigella sonnei*. We focus on the emergence of AMR, but EMERGENe has significant potential to be used in a much wider research and surveillance context to robustly characterize and compare trait behaviour in bacterial populations.

## Methods

### EMERGENe model overview

We represent the phylogenetic tree as a directed acyclic graph (12) defined as:

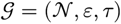

where:

*N* is a set of 2*n*−1 nodes, representing sampled and ancestral lineages, *ε* ⊆ {(*u, v* | *u, v* ∈ *N*} is a set of 2*n*−2 directed edges, with each edge (*u, v*) indicating that *u* is the parent of *v*,

τ : *N* → ℝ_≥0_ assigns each node a time value, with τ(*u*) < τ(*v*) for (*u, v*) ∈ *ε*.

### Detect Phyletic Events through Ancestral State Reconstruction

We apply ancestral state reconstruction of binary traits through stochastic character mapping using the "make.simmap" function from the R package "phytools" (27). The function was set using all-rates-different model (ARD), as we assume that the transition rates between the states i.e. gain of resistance may occur at different paces, therefore the gain and loss of these traits are unlikely to be equal in this context. In addition, we set the prior of the root node was estimated via the stationery distribution as pi*Q=0. After estimating the transition-rate matrix Q, we generated 100 stochastic mappings of character evolution, then summary statistics were obtained.

Then, we consider each node *u* ∈ *N* to be associated with a binary state based on the internal state node probability (only nodes with minimum node probability of 0.8 are considered):

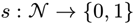

where 0 and 1 represent two discrete character states of a trait (See Figure 2). For an internal node *u*, we denote its child as (*u*) = {*v*_1_, *v*_2_}.

**Figure 2.**
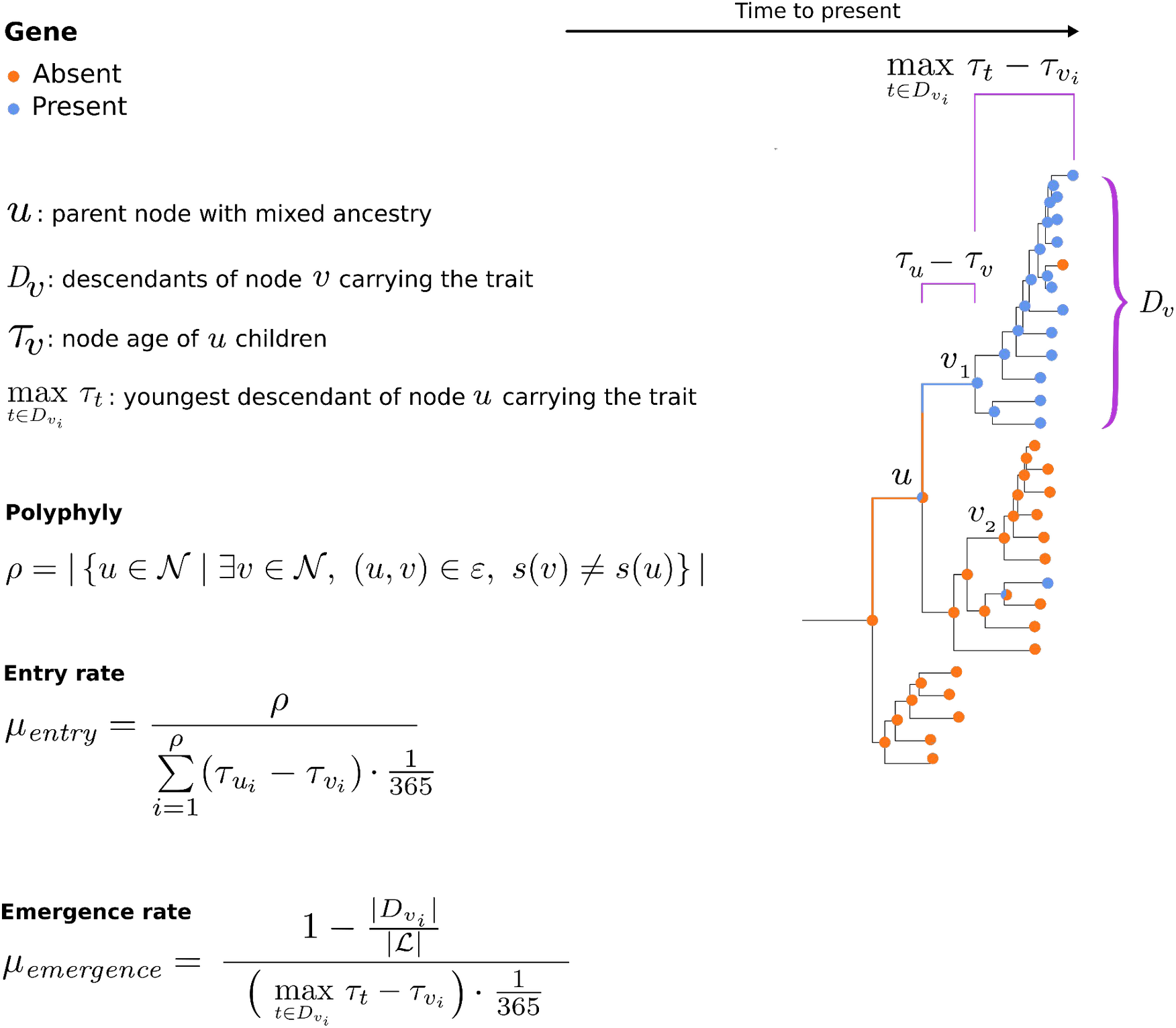
An overview of EMERGENe. Parameters are shown on an example phylogenetic tree (right) with a single trait overlaid as presence/absence according to the inlaid colour key defined upper left (or partial state probability indicated by multicolor nodes). Output metrics and equations are shown lower left, with further detail on mathematical parameters available in Methods.

### Phyletic events - Introductions

Once that the history of a trait is reconstructed, the number of introductions of the trait across the population are detected, intended when the state of an ancestral node shifts the state i.e. trait acquisition, as previously shown (27,28). We define the set of nodes that undergo a state transition from the parent node to at least one child as:

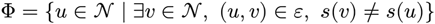

The total number of transitions, referred to here as phyletic events, is given by:

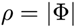

The number of phyletic events define the nature of the phenotypic trait, monophyletic if this is observed once in the tree or polyphyletic when observed multiple times i.e. homoplasmic.

### Entry rate - Trait introduction

We calculate the entry rate as:

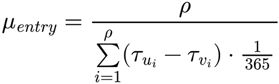

Where τ*_u_i__* − τ*_v_i__* is the branch distance between the parent node and its child with opposite state i.e. branch duration associated with the trait introduction, where a transition between demes occurs. Entry rate is expressed by phyletic events per branch-year

### Emergence Rate — Post-Introduction Trait Propagation

Following each introduction event, defined as a transition from a parent node *u_i_* to a child node *v_i_* with *s*(*v_i_*) ≠ *s*(*u_i_*), we quantify the Emergence rate as the degree to which the newly acquired trait spreads during the early phase of its presence in the population. For each introduction event at node *v_i_*, we define

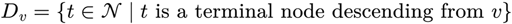

as the number of such descendant terminal nodes. Let

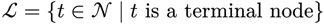

be the total number of terminal nodes in the phylogeny, and the propagation time is given by:

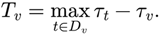

The emergence rate is then:

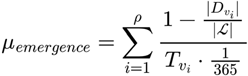

This metric increases when (i) the trait remains rare (small descendant clade), and (ii) the branch length distance is short i.e. lineages accumulate rapidly in time. Conversely, the metric decreases when the trait is already highly prevalent, reflecting the loss of early-stage emergence information. The emergence rate therefore captures the time of initial post-introduction emergence, rather than long-term trait prevalence.

Emergence rate is expressed as cases per branch-year, as we divide (including the Entry rate) the phylogenetic tree branch lengths on the number of days within a year (See Figure 2).

#### Trait emergence simulations

To test EMERGENe, we performed different phylogenetic simulations using ReMASTER v2.7.7 (29). Four different populations (i.e. demes) were included following a structured coalescent trajectory, which accounted for migration events across demes (i.e. in the case of bacterial traits horizontal gene transfer arising from e.g. plasmid transfer, recombination) to make our case study more realistic. Briefly, we set a *background population – constant growth* population with a single bottleneck event reducing the effective population size (Ne=50 to Ne=30, Green, Figure 3B). This might represent a background antimicrobial susceptible population which is being replaced by the emergence of multiple AMR clades. From this population, two populations representing the acquisition of two distinct traits which grew exponentially, but at different rates. These were a population with *ancestral expansion – full capacity* (Ne=60, Growth rate=0.5, Yellow) and another with *a recent expansion – full capacity* (Ne=20, Growth rate = 1, Blue, Figure 3B). Hence, the first (Yellow) reached maximum capacity prior to the second (Blue), which had more recent higher growth rate, these may, for example, represent an older AMR profile (e.g. tetracyclines) and a more recently emerged one (e.g. ampicillin) respectively. We added a final population *a recent expansion – still growing* (Ne=10, Growth rate=3, Purple) with rapid growth that emerged from the *ancestral expansion – full capacity* population (i.e. Purple emerged from Yellow, figure 3B). This represents rapidly emerging traits in need of early identification such as AMR against the most recent antimicrobials (e.g. third generation cephalosporins). Population coalescent and size parameters were selected to be consistent with previous work (9,29).

**Figure 3.**
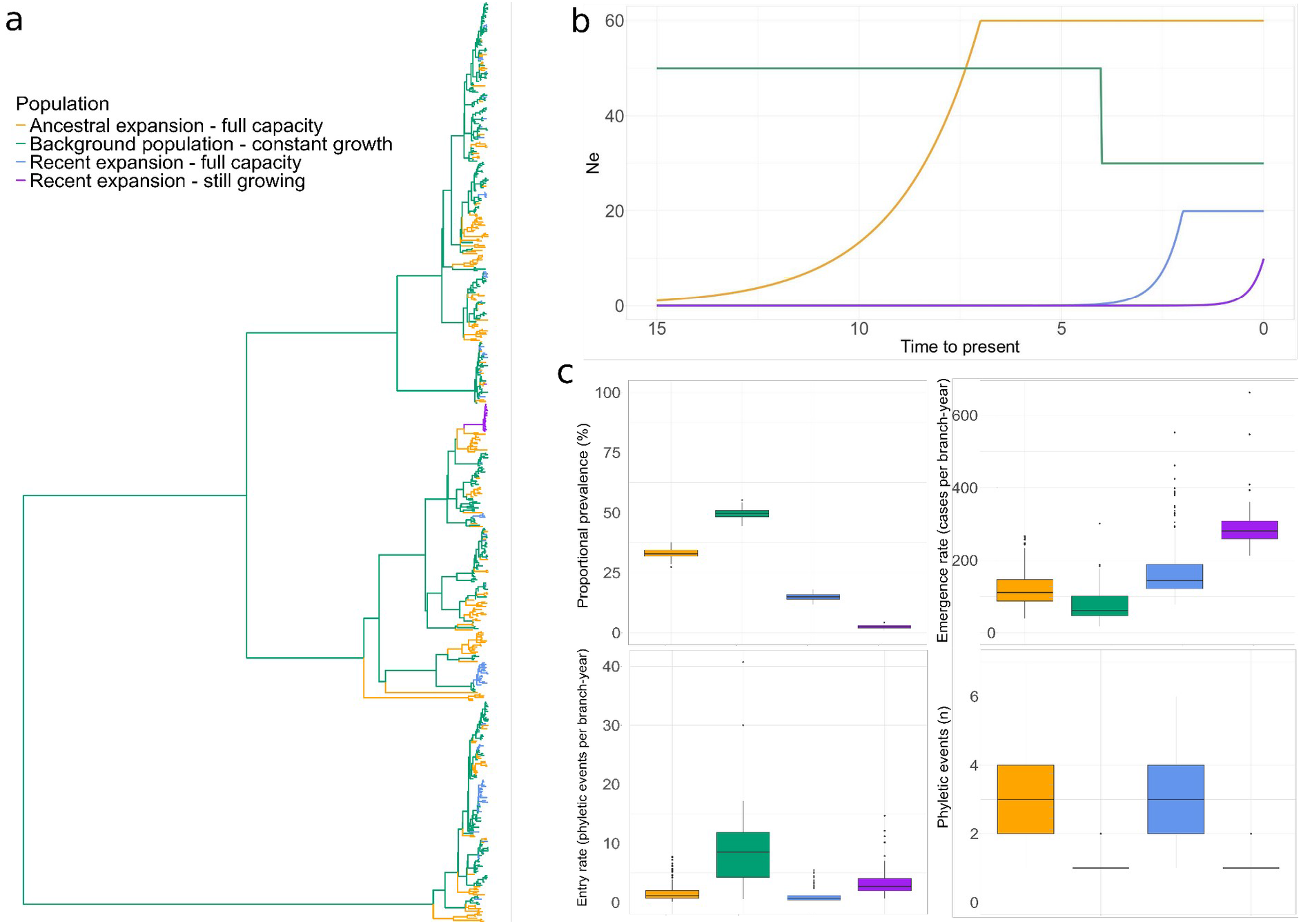
The efficacy of EMERGENe in capturing trait behaviours in dynamic populations. Leftmost (**A**) is a single example phylogenetic tree and trait distribution resulting from a ReMASTER simulation of four populations whose effective population sizes varied over time (shown in **B**). The tree tips are colored according to population type. The bottom right panel (**C**) shows boxplots (coloured by trait/population), comparing the median distribution of measures from 100 ReMASTER simulations. These include the Proportional presence (i.e. standardly used ‘prevalence’) of the traits compared with the EMERGENe metrics; Emergence rate, Entry rate and Phyletic events over the simulations.

We ran 100 phylogenetic simulations of these populations (with a mean of 603 isolates) and then mapped the four different traits (represented by each deme/population) across each phylogeny (which we assumed were carried by all individuals consistent with the trait ancestral state reconstruction in EMERGENe). We then simulated gene transfer events between different lineages by setting migration events to arise at different rates during the transition events of the population with *recent expansion – still growing* from the *ancestral expansion - full capacity,* and *recent expansion – full capacity* from the *background* respectively. And finally, ran EMERGENe on each simulation, for each trait,to compare trait EMERGENe metrics and proportional presence (i.e. currently reported ‘prevalence’). Here, we reported only phyletic events with five or more terminal tips to reduce possible overestimation of Emergence rate due to small, negligible clonal effects (30) that can arise stochastically from the simulations. We also restricted the analysis to parent nodes where the state probability was >=0.8. Collectively, this ensured a reliable assignment of ancestral nodes and robust inference of quantitative EMERGENe variables. Notably however, users can tune these parameters according to their needs.

#### Comparing traits using EMERGENe in real world surveillance data

As EMERGENe requires a timed tree, we used a large genomic surveillance dataset of the globally important diarrhoeal pathogen, *Shigella sonnei*, generated by the United Kingdom Health Security Agency. Briefly, this data comprises 3,745 strains isolated from 2012 to 2020 in UK (Bioproject number: PRJNA248792) and comprises a maximum likelihood phylogenetic tree (from (31) and molecular clock analysis using Bactdating(32), setting a default clock rate with Gamma prior (0.001, 1000) and 1x10^8^ Markov Chain Monte Carlo iterations as previously described (33). To examine trait behaviour, we generated AMR and virulence gene presence and absence over the surveillance dataset using AMRFinderPLUS v3.11.2 (34) with the following filters: coverage 100% and >99% identity; and generated a genome presence/absence matrix. To compare EMERGENe rates related to point mutations, fluoroquinolone resistance associated mutations in *gyrA* and *parC* were also included.

#### Detailed analyses to simulate public health use

Owing to their clinical and public health relevance, good genotypic and phenotypic concordance, and our understanding of the recent dynamics of these resistance profiles, we additionally looked at the temporal dynamics of three specific predicted AMR phenotypes: azithromycin (AZM), ciprofloxacin (CIP), ceftriaxone (CRO), and a recently emerging phage–plasmid pWPMR2, with initially cryptic function (i.e. with no known fitness advantage) but latterly associated with AMR evolution (31). Specifically, AZM was defined by the presence of *ermB* and/or *mph(A)*; CIP by the presence of triple mutations consisting of S83L and D87G/N in *gyrA* together with S80I in *parC;* CRO by the presence of *blaCTX-M* variants; and pWPMR2 by mapping against the phage–plasmid sequence. All genetic features were encoded as binary traits.

To assess temporal changes in evolutionary rates (i.e. simulating real time surveillance), the dataset was partitioned into a series of cumulative yearly windows, with a fixed starting point from 2012 to successive endpoint years across the full temporal span of the dataset.

_Let_ *Y* ={ *y*_1_,… *, y_n_* }*, y*_1_ = *y_min_, y_i_*_+1_= *y_i_* +1 *, i*=1… *, n−*1 . _For each year_ *t* ∈ *Y* _, with_ *X_t_* denoting the set of isolates sampled in year *t .* For each retrospective iteration *i*=1, …*, n,* with endpoint *y*= *y_i_* = *y_min_*+*i −*1, the cumulative dataset is defined by *D_yi_*=_{_∃*t* ∈ _{_*y_min_,* … *, y_i_* _}_*, x* ∈ *X_t_* _}_

Starting from the main phylogenetic tree, for each time window *y* > *y_min_*, the cumulative dataset was defined as the isolates sampled from the starting year to the next endpoint, pruning the tips beyond the time window from the main tree. Then, for each time window, a time-specific subtree representing the information available at that retrospective time point was generated. Evolutionary rate estimates for each resistance determinant were then calculated independently on each subtree. At times to reduce noise (see Results), two filters were applied to the minimum number of tips needed to define Phyletic events that contributed to trait Entry and Emergence rate estimation including two- and five- tip thresholds. These thresholds allowed us to assess the sensitivity of EMERGENe (e.g. how many isolates were necessary to spot an event at the early stage) to detect emerging traits while also accounting for stochasticity in the data and tree reconstruction.

## Results

### EMERGENe model overview

EMERGENe takes as input user provided time-scaled phylogeny (newick format) and trait presence-absence information in table format. Then, the software employs ancestral state reconstruction (ASR) through stochastic character mapping following an all-rates-different model, allowing multiple state changes across tree branches while accounting for evolutionary time. Uncertainty of character states on tree nodes are also accounted for through marginal posterior probabilities in the summary of sampled stochastic maps. Pairing these ASR states with phylogenetic distance and tree topology, EMERGENe then estimates three novel metrics for each binary trait in the bacterial population. Specifically, a summary table is generated for three primary outcome metrics (per binary trait). These are, Phyletic events (n) which approximate the number of independent trait acquisitions; the Entry rate (phyletic event/branch year), and Emergence rate (in cases/branch year). Additional granularity on the latter two traits is provided for each trait Phyletic event per internal node (i.e. with the ancestrally reconstructed shift in trait acquisition) is also provided, including: the ASR node state probability; the number of descendant internal and terminal tips (i.e. cases); and node age. The code to generate the output tables, and R code to generate plots from the main output table are available at https://github.com/gbatbiff/EMERGENe.

To expand on the novel Entry and Emergence rate metrics, once a Phyletic event on an internal node is detected (e.g. when a trait is gained resulting in mixed ancestry, *u* nodes, Figure 2), Entry and Emergence rates are calculated.

The Entry rate represents the time that passed during a lineage’s existence to a change in state in descendant nodes (e.g. from absent to present, see transitionary *v* nodes and *t _u_−t_v_* in Figure 2). The Entry rate represents the time required for the trait to be introduced in the population for example through horizontal gene transfer or recombination. Hence, a high Entry rate would indicate a slowly emergent trait acquisition such as a highly costly AMR plasmid which needs to ameliorate cost before expansion is observed, or entry of the bacteria to a new ecological niche with strong negative selection driving trait acquisition (i.e. where variants that arise from genetic drift are removed and stabilising selection fixes advantageous traits). The Emergence rate captures the magnitude of the subsequent expansion after the trait is acquired (i.e. the time over which a trait is present after its introduction in the parent node and the associated expansion of the lineage as a function of the number of terminal tips bearing the trait, Figure 2). For a typical epidemiological dataset in an AMR surveillance context, this would be akin to the number of cases caused by a new AMR strain over time since its most recent common ancestor. In this scenario, a high Emergence rate is an indication of how rapidly a clone having acquired the trait can expand, whereas a low Emergence rate might indicate acquisition of a necessary, but highly costly AMR phenotype.

### Testing EMERGENe on phylogenetic simulations

To test the utility of our method for detecting trait emergence beyond proportional presence data, we performed phylogenetic simulations including four populations (simulating different traits) with distinct evolutionary trajectories (Methods, example phylogeny Figure 3A). In this analysis, a population with constant evolution and a bottleneck to reduce the effective population size was set to simulate a background population in which traits emerge (Green, Figure 3B). Three populations emerged from this background to simulate trait acquisition (e.g. through horizontal gene transfer) and spread among the population. Two polyphyletic traits emerged to fixation in the population (i.e. the populations reached full capacity) with one being ancestral and slower growing (Yellow, Figure 3B) than another more recent population with a faster growth rate but lower overall population size (Blue, Figure 3B). A final monophyletic population had a comparatively small population size but had not reached fixation and was still rapidly growing (Purple, Figure 3B). This final population represents a major gap in pathogen genomic surveillance where small, but rapidly growing populations, are not highlighted as issues in proportional presence data. Hence, in our simulations, we captured the assemblage of challenges in AMR surveillance using routine genomic epidemiology, including, critically, the limited resolution of proportional presence.

When evaluating EMERGENe on the simulations, we compared the results of our method with proportional presence (Figure 3C). This revealed that proportional presence was biased toward older populations reflecting historical presence, rather than highlighting novel, rapidly emerging traits. This was distinct from the Emergence rate variable which more closely mirrored the growth rates of the populations. Specifically, the aggregated Emergence rate of *recent expansion – still growing* (purple) and *recent expansion – full capacity* (blue) were 292 and 164 cases/branch year respectively, with *ancestral expansion – full capacity* (yellow) having 121 cases/branch year, consistent with the lower growth rate set in the simulation. The *background – constant growth* (green) exhibited an average of 86 cases/branch year; significantly lower than the others, as expected, and in stark contrast to its high prevalence value.

When considering the other EMERGENe variables, these were consistent with our population simulations and offered an orthogonal view on traits Emergence rate. Specifically, regarding Phyletic events, our method correctly recovered the convergent evolution permitted in the populations with a median of three acquisitions (i.e. Phyletic events n=3) being found for the *recent expansion – full capacity* population, and only one in the *recent expansion – still growing* and *background population – constant growth* populations (Figure 3C). The Entry rate was also estimated correctly, being higher (i.e. arising from a slower introduction, see long branch preceding the Purple population expansion in Figure 3A), about four times smaller in the *recent expansion – full capacity* (Blue). Collectively, this demonstrates that EMERGENe variables are well suited to detecting rapidly expanding traits in bacterial populations while also offering improved granularity and an orthogonal approach to reporting proportional presence alone.

Two findings were further explored in the simulation data. These included the detection of Phyletic events for the *background population - constant growth* which was not expanding. These likely arose from phylogenetically proximate phyletic and/or migration events of the other traits in our simulations, but could also occur in genuine surveillance data consequent to natural biological processes (e.g. plasmid loss). These artefacts were infrequent, with only 1 to 2 Phyletic events (mean=1.15, SD=0.37) of the background population across only 19 of the 100 simulations. Although we do not consider this a major issue (as EMERGENe primarily functions to examine trait acquisition from background), this prompted us to determine the impact of Phyletic event size (i.e. number of descendant terminal tips) on the Emergence rate. This revealed that a lower number of tips was associated with a higher Emergence rate (Supplementary Figure 1). This is not unexpected as Emergence rate is calculated mainly based on branch lengths between nodes (i.e. coalescent times) and the number of descendant lineages, so the rate increases as the number of terminal tips decrease.

### Application of EMERGENe to study AMR emergence

Having demonstrated the efficacy of EMERGENe in simulated data, we tested EMERGENe performance on real genomic surveillance data of *Shigella sonnei* (n=3,745 genomes) from the United Kingdom sampled between 2008 and 2020 (31). A time-scaled phylogenetic tree and the AMR and virulence gene content were used as input (from (31,33) and EMERGENe metrics were inferred (Figure 4). Here, we compare the Phyletic events, Entry and Emergence rates. These were inferred with a minimum threshold of two terminal tip descendants, which is consistent with our simulation findings, previous work on migration rates (35), and the public health aim of picking up rapidly emerging traits with high sensitivity before a large number of cases occur. These results were also consistent with the stricter filter of a five-tip minimum threshold (Supplementary Figure 2).

**Figure 4.**
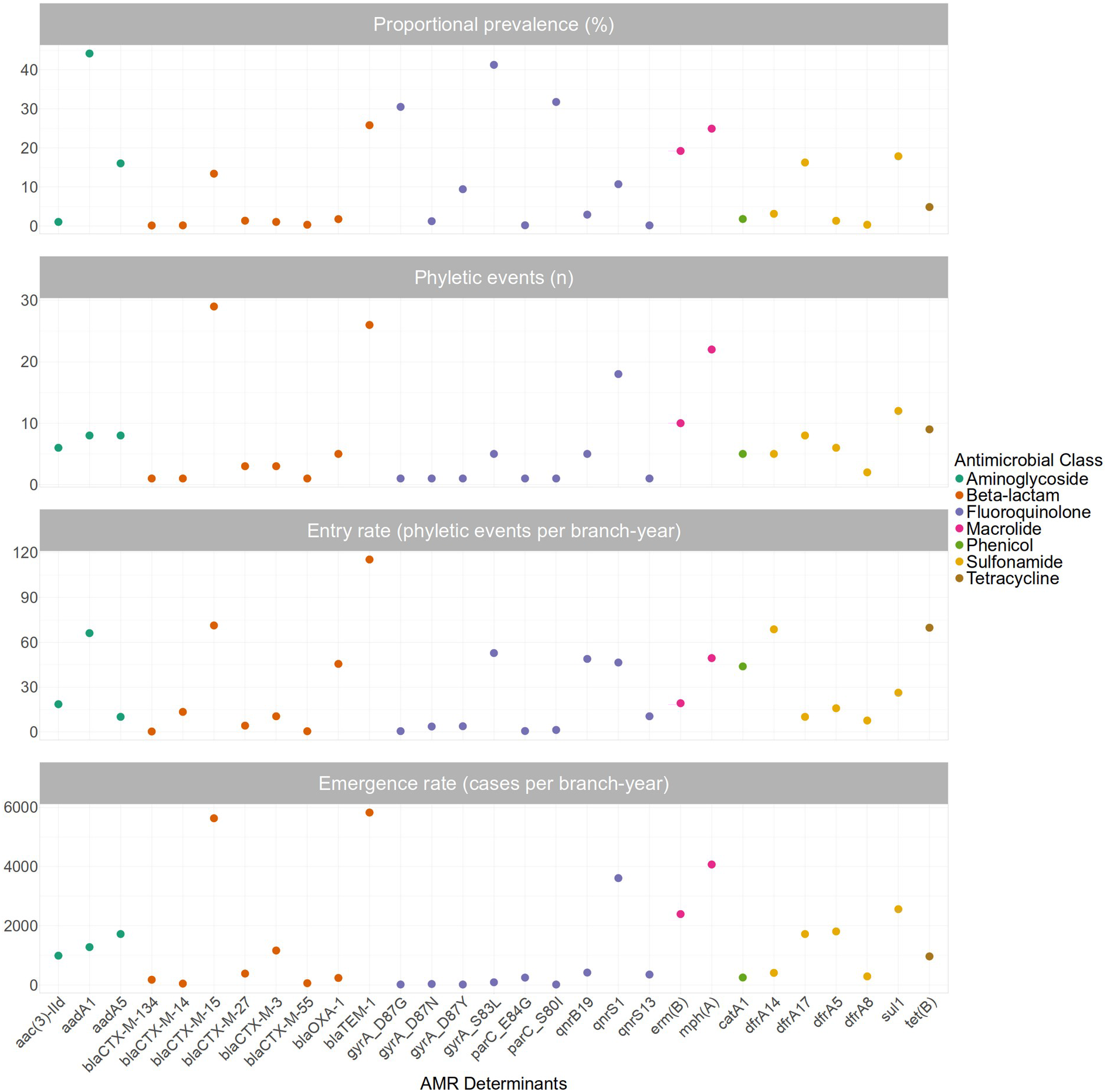
EMERGENe extends AMR characterisation beyond proportional presence. The graphs show proportional presence (upper) and the variables obtained using EMERGENe (lower three) for AMR and virulence genes from *S. sonnei* surveillance data (n=3,745). Starting from the upper panel: i) Proportional presence of AMR genes ii) Polyphyly iii) Entry rate iv) Emergence rate.

As anticipated, EMERGENe was effective at discriminating the emergence of different AMR genes in a manner distinct from proportional presence and highly consistent with reported epidemiological trends in *S. sonnei* populations. Specifically for example, the aminoglycoside resistance encoding *aadA1* gene was found at high prevalence (44% of isolates) but had comparably low Emergence rates and number of Phyletic events, consistent with this antimicrobial not being in current clinical use so exerting little current selection pressure for the accessory genome (Figure 4). Contrastingly however, genes related to azithromycin (*mphA*) and third-generation cephalosporin resistance (*bla*_CTX-M-15_), well characterised clinical selection pressures in this setting (33) had lower prevalences (24.9% and 13.4% respectively) than *aadA1*, but much higher Emergence rates and Phyletic event counts (Figure 4). This is consistent with a reported decade of emerging azithromycin resistance and, the recent dramatic emergence of resistance against extended-spectrum beta lactamase in clinical strains, most of which took place after 2020 (the end of this sampling frame)(36). Hence, EMERGENe is effective for highlighting genetic traits that are rapidly emerging in bacterial populations.

We also found that EMERGENe detected the undescribed but intuitive rapid emergence of other traits. For example, another highly clinically relevant antimicrobial class for *Shigella* is fluoroquinolones, and while EMERGENe highlighted the rapid emergence of the horizontally transmissible *qnrS1* gene, the metrics of this gene were distinct from the vertically inherited fluoroquinolone resistance encoded by chromosomally encoded mutations in *gyrA* and *parC*. This highlights the difference in emergence potential of horizontally and vertically inherited AMR traits and indicates a rapid, but undescribed spreading of the *qnrS1* genes in the bacterial population. In addition, we observed high rates of emergence, including a high number of phyletic events for the disinfectant (specifically, quaternary ammonium compounds) resistance protein *qacEdelta1*, suggesting EMERGENe can be used to spot overlooked (i.e. low prevalence and/or poor functional understanding) genes that are rapidly disseminating in bacterial populations. Notably, although focused on comparing AMR gene epidemiology across our dataset, EMERGENe can be used to measure the emergence of any bacterial trait including phenotypic AMR (either empirically measured or genotypically inferred). To test this, we genotypically inferred AMR phenotypes and aggregated these to antimicrobial class and found patterns consistent with gene level patterns (Figure 4; Supplementary Figure 2).

### EMERGENe for monitoring AMR trends in surveillance data

To evaluate the surveillance potential of EMERGENe as a tool for identifying rapidly emerging traits, we then conducted a temporal simulation analysis using the surveillance data. We focused this analysis on four traits whose dynamics are well characterised in the *S. sonnei* population, genotypically predicted Azithromycin (AZM), Ciprofloxacin (CIP), and Ceftriaxone (CRO) resistance (see methods) and a recently discovered phage plasmid pWPMR2 (31), which promotes the evolution of AMR. Initially, we calculated the overall rates for each of these traits considering the two filters, to have a proxy of their behaviour over the whole sampling period based on the minimum threshold used (Supplementary Figure 3). Here, we observed how the CRO resistance experienced multiple but slow introductions i.e. high entry rate, but then it rapidly spread across the population. CIP originated from one single monophyletic event before reaching high capacity in the population, followed by a broad but slow dissemination. Whilst phage plasmid pWPMR2 exhibits a similar evolutionary pattern of AZM, being known to be evolutionary successful in *S. sonnei*, supporting the potential of EMERGENe to detect traits whose trajectories can be epidemiologically relevant.

To explore the temporal performance of EMERGENe, we tested cumulative versions of the dataset run to the end of individual years in the sampling frame (i.e. where 2018 includes all data from 2012 to 2018). This was done by generating cumulative sub-trees over the years on which we inferred EMERGENe metrics. We then compared trends in these variables with trends in proportional presence as a baseline for current AMR surveillance practice, applying the two filters on minimum number of tips (Figure 5; Supplementary Figure 4). Our temporal analysis revealed a universal increase in proportional presence across each of the traits, to which EMERGENe was able to bring greater granularity. Specifically for example, the number of introductions of each trait (i.e. Phyletic events) were increasing over time in all cases except for CIP, which experienced a large and monophyletic acquisition of the trait (Figure 5; Supplementary Figure 4), and see (31) underpinned by its chromosomal location and vertical inheritance pattern. Despite having a similarly increasing proportional presence to other AMR measures, CIP had significantly lower EMERGENe variables, highlighting the suitability of EMERGENe for identifying rapidly emerging transmissible traits.

**Figure 5.**
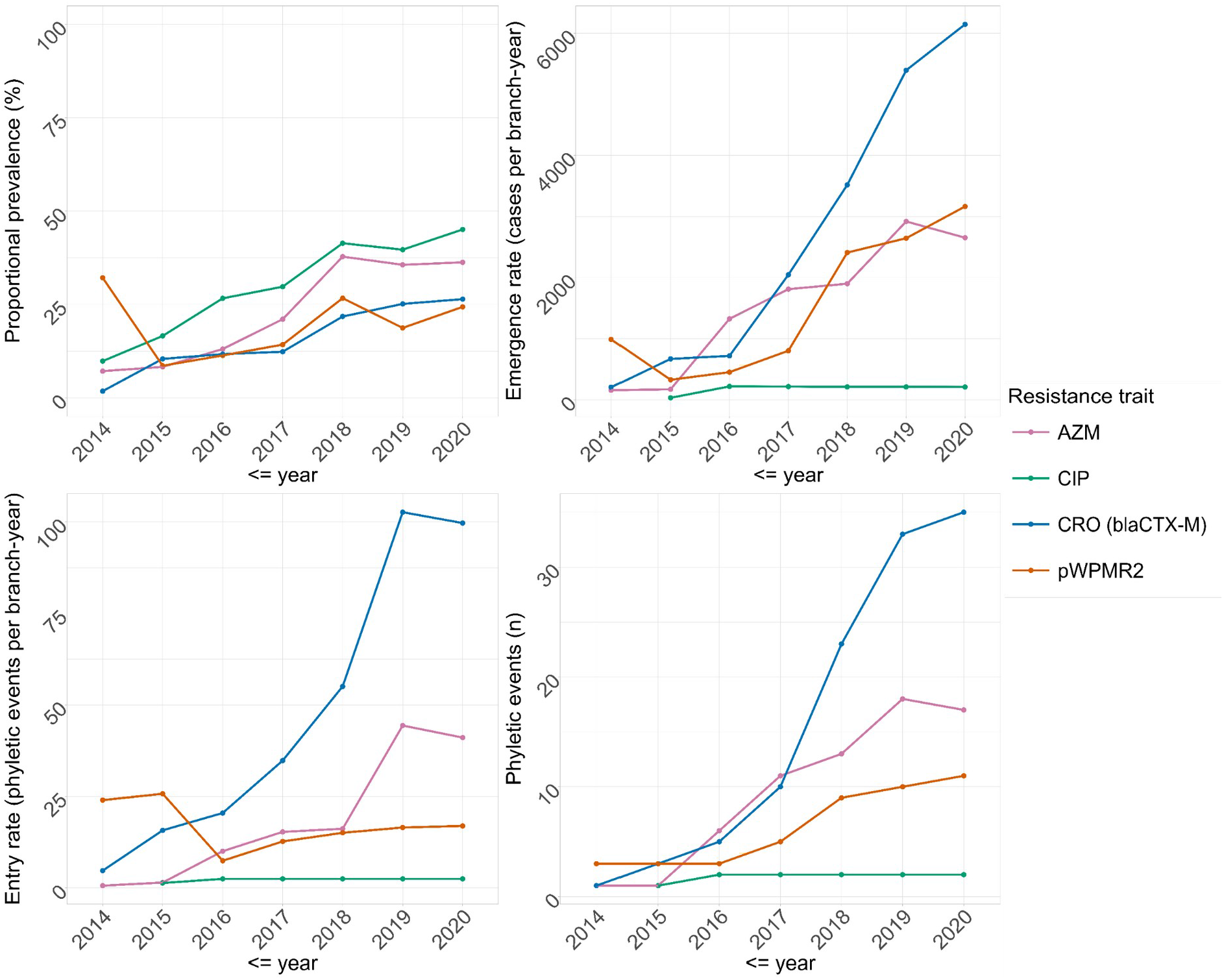
Temporal trends in EMERGENe metrics relative to proportional presence. Time series of *Shigella sonnei* trait emergence. Plots show trends in i) Proportional presence (upper left) ii) Emergence rate (upper right) iii) Entry rate (lower left) and iv) Polyphyly (lower right). Data for four relevant AMR traits are shown coloured according to the inlaid key. NB: As there were much smaller numbers of samples in 2012 and 2013, EMERGENe variables were not inferred for these years.

This temporal analysis also revealed a comparatively rapid emergence of AMR against third generation cephalosporin antimicrobials (CRO, Figure 5). Specifically, while AZM emergence increased steadily in the population it plateaued in 2019 when treatment guidelines changes moved away from AZM and drove down fitness of AZM resistant strains (33). Similarly, the phage plasmid pWPMR2 exhibited a comparable emerging trend of AZM, which was still ongoing in 2020, indicating how EMERGENe could be used not only to trace already known traits, but also for novel emerging properties of epidemiological interest. By contrast, CRO emergence indicators from EMERGENe remained on an upward trajectory and were predictive of the subsequent large-scale outbreaks of CRO-resistant *S. sonnei* after the sampling frame of this window. Notably, the extended information on individual phyletic events could also be used to gain an overview of the temporal epidemiological patterns of trait emergence in the population (i.e. which clades and how many of them and when, Supplementary Figure 5).

## Discussion

In this work, we present EMERGENe, a phylodynamic method based on ancestral state reconstruction that provides a robust comparison of epidemiologically relevant rates related to the evolutionary dynamics of binary traits across a given population. We tested our method both on simulated and real data, showing its reliability to estimate the emergence of transmissible traits, such as AMR. EMERGENe metrics accurately recovered population dynamics across simulations and outperformed standard proportional presence for capturing recent evolutionary dynamics, including over simulated time courses.

Our temporal analysis on the national surveillance dataset was designed to resemble its potential application in routine genomic surveillance. This recaptured recent evolutionary dynamics, with CRO and pWPMR2 emerging more rapidly in terms of both Phyletic events and Emergence rates, followed by AZM resistance and the established CIP resistance. This is consistent with the described spread of CIP resistance into the UK between 2008 and 2014 (37) followed by the subsequent acquisition of transmissible AZM and CRO resistance (36). EMERGENe recaptured these dynamics by spotting emergence of CIP at the early stage of its spread, followed by a more stable trend until 2020, indicating that CIP resistance progressively fixed and reached maximum capacity in the population. Contrastingly, the number of Phyletic events and Emergence rate were significantly higher, for CRO resistance (not evident in the temporal trends in proportional presence) which was predictive of its subsequent rapid rise since this dataset ended in 2020 (38). Furthermore, EMERGENe was also able to point to previously unreported emerging genes (e.g. *qnrS1*) and those with cryptic function (pWPMR2). Hence, our simulated surveillance exercise highlighted the utility of EMERGENe as a potential public health tool for epidemiologically relevant and unknown traits.

The application of EMERGENe also opens several exciting research avenues for the evolutionary dynamics of transmissible traits in bacterial populations. For example, in those cases where the Emergence rate was comparatively high (e.g. *bla*_CTX-M-15,_ *bla*_TEM_, and *mphA*) we also observed comparatively high Entry rates. Ecologically, this means that slow entry/migration of the trait was followed by rapid expansion of the trait in the population. It is likely that this pattern captures pulsed evolution, such as the punctuated equilibrium initially theorised by Gould and Eldredge (39), and recently observed in bacteria across the tree of life (40), where long periods of stasis occur at a population stall or saddle point are then followed by sudden burst of evolution and rapid diversification (41). In the field of infectious disease epidemiology, this could be thought of as a variant fitness valley, requiring adaptation before emergence in a novel host. Hence, EMERGENe offers exciting potential to more broadly develop and test hypotheses regarding the impact of accessory genome changes on evolution.

Although not focused on here, EMERGENe returns detailed granular information about the age and magnitude of all phyletic events associated with a given trait, which can serve as an important descriptive portrait of the natural history of trait emergence (Supplementary Figure 5). Specifically, this portrait can reveal when distinct trait-bearing clades occurred and how many cases resulted. This can be important information for exploring EMERGENe results qualitatively. For example, in the case of CIP resistance, it becomes clear that this is highly present in the population due to a single ancestral phyletic event. This large, monophyletic expansion is expected given our strict three-chromosomal mutation encoding of the phenotype (though notably higher signals of convergence would have been observed with a relaxed single mutation encoding as in gyrA/*parC* in Figure 5; Supplementary Figure 3; Supplementary Figure 5) but is in stark contrast to the three other plasmid-encoded traits (i.e. AZM, CRO, pWPMR2) which show a trend of many acquisitions of high Emergence rate later in time. This highlights the need to consider the number of Phyletic events alongside EMERGENe variables and suggests EMERGENe may aid in the differentiation of vertically and horizontally encoded traits, such as AMR.

Although showing great promise, it is important to note several limitations and areas for future development of EMERGENe. Firstly, the sensitivity of EMERGENe to different tree reconstruction parameters and the influence of uncertainty has not been assessed so users need to have high confidence in the provided time tree. Secondly, owing to the intuitive inverse correlation between Emergence rate and clade size, there is a need to tune the minimum tips sensitivity threshold. We suggest users do this by exploring correlations within their own datasets and by assessing performance of multiple minimal tip thresholds. Furthermore, as with any association evolutionary change with epidemiological events there is a need to consider genetic linkage so users should be cautious to explore co-occurrence and synteny among traits when interpreting EMERGENe metrics. Additionally, EMERGENe only accounts for gain events, thus not considering trait acquisition if its proportional presence in the analysed population is too high (i.e. the trait is ≥50% proportional presence) though this can be tuned by the user. Finally, EMERGENe provides absolute rates that can scale linearly depending on the number of phyletic events, making it difficult to compare rates across different sampling collections where bacterial populations size and AMR dynamics can vary greatly. Hence, its main utility should be observed when analysing and comparing trajectories of traits within a given population.

Despite these caveats, we demonstrate here that EMERGENe is a promising and user-friendly tool that can be used to broadly identify and compare emergence of traits in bacterial populations. The consequent ability to formally quantify, particularly transmissible, trait emergence opens various avenues of further research and applications in public health. Specifically, early identification of traits of epidemiological interest (such as AMR) would facilitate onward timely and targeted interventions to prevent and mitigate spread (e.g. outbreak prioritisation and engagement with clinical treatment advisory bodies). EMERGENe can be easily implemented as it takes standard inputs format and provides clear, interpretable summary results and represents an early and unique tool for studying accessory genome dynamics across bacterial population structures.

## Data Availability

All data produced in the present work are contained in the manuscript

https://doi.org/10.1016/S1473-3099(26)00227-6

