## Supplementary_Material for "Estimating emergence rates of epidemiologically relevant traits in bacteria with EMERGENe"

### Supplementary information

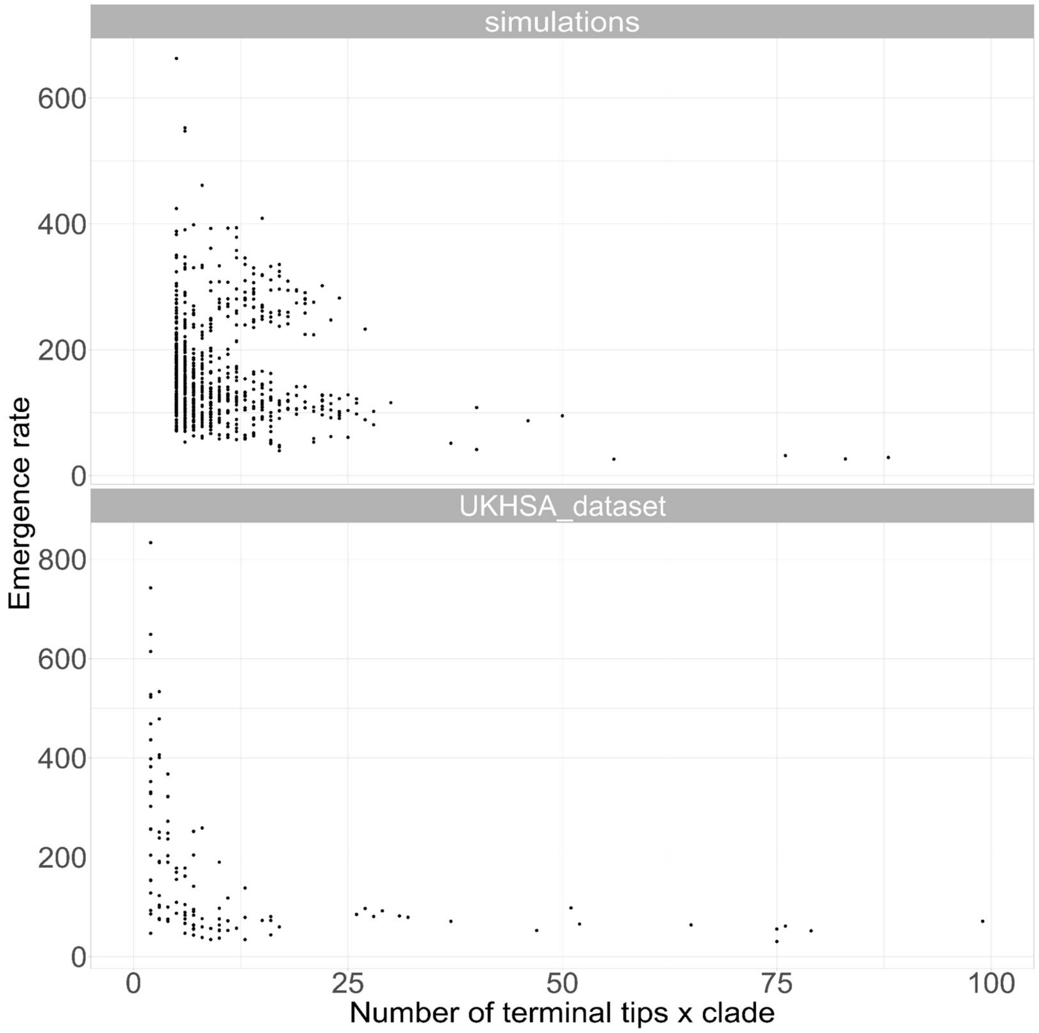


**Supplementary Figure 1. Inferred EMERGENe Emergence rate across simulations (top) and UKHSA dataset (bottom)  by lineage size.** The Emergence rate of the polyphyletic events is shown as a function of the number of terminal tips (xlim set to 100 for visualiszation) descending from each event.

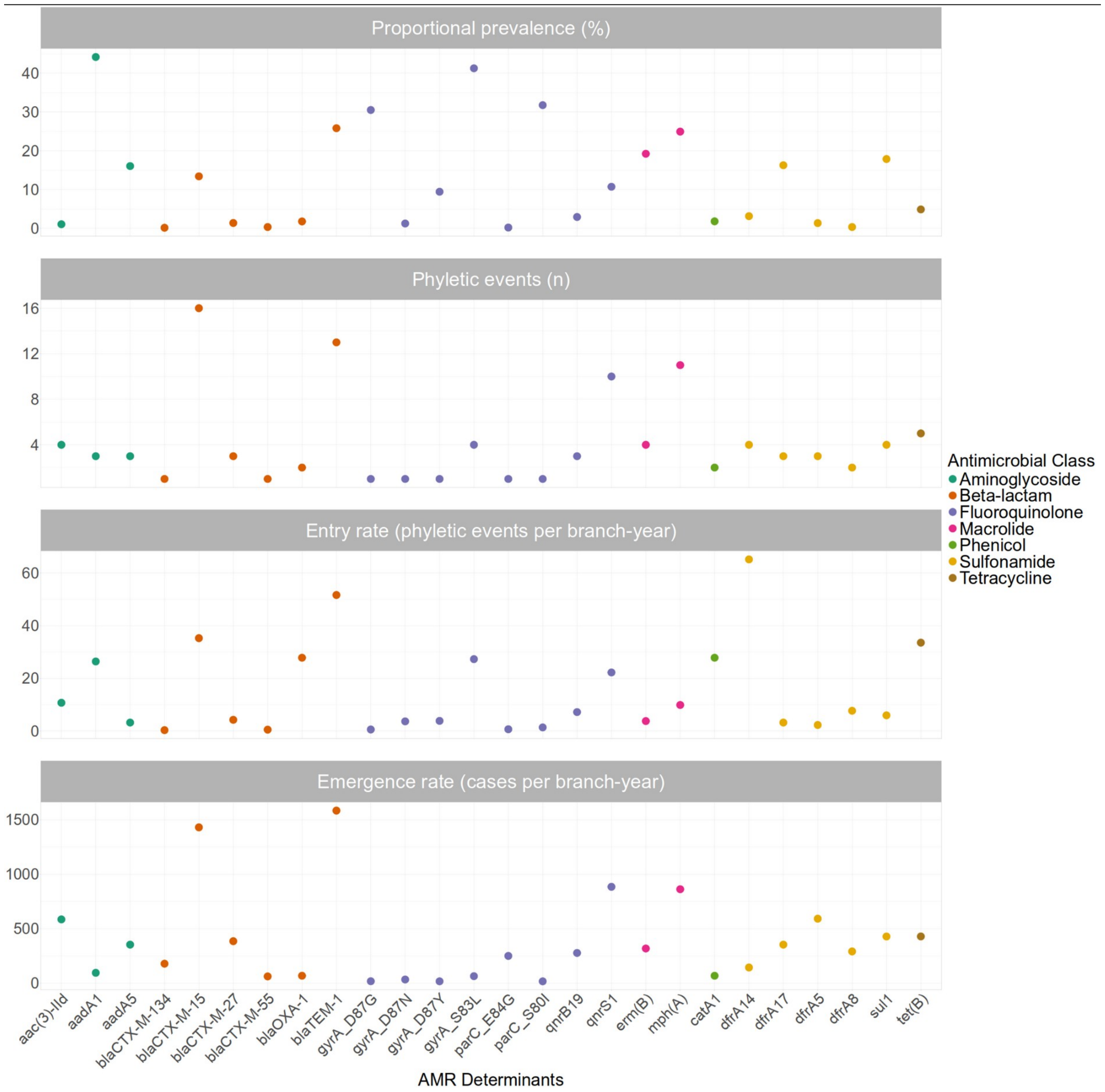


**Supplementary Figure 2.** Output variables obtained from using EMERGENe on the AMR and virulence genes from 3,745 *Shigella sonnei* from routine surveillance at UKHSA, including clades with a minimum of five descendants from the trait ancestral node. The panels show the standard commonly reported proportional presence of the genes in the dataset adjacent to EMERGENe variables. Starting from the upper panel these are: i) Gene proportional presence (prevalence) in the dataset ii) EMERGENe inferred Polyphyly iii) Entry rate iv) Emergence rate, the time and magnitude of trait spread across the population once introduced.

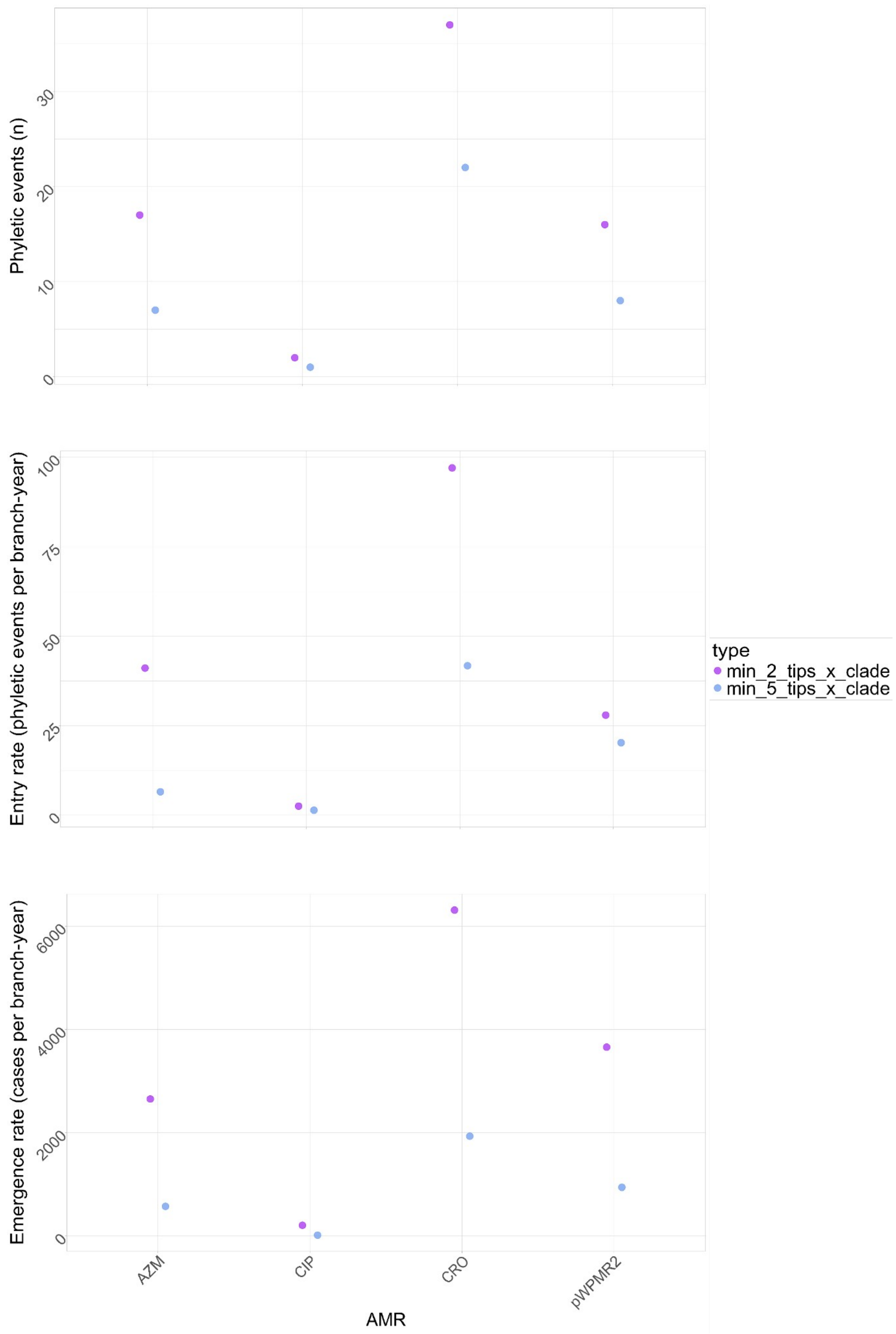


**Supplementary Figure 3. EMERGENe estimates for four major AMR phenotypes including the impact of tip thresholds.** EMERGENe variable values are shown for genotypically inferred traits of azithromycin (AZM), ciprofloxacin (CIP), and ceftriaxone (CRO) resistance, and the presence of the pWPMR2 phage plasmid. Variables are shown as measured for both unfiltered (i.e. minimum of two tips, purple) and filtered (blue, minimum of five tips per clade) analyses.

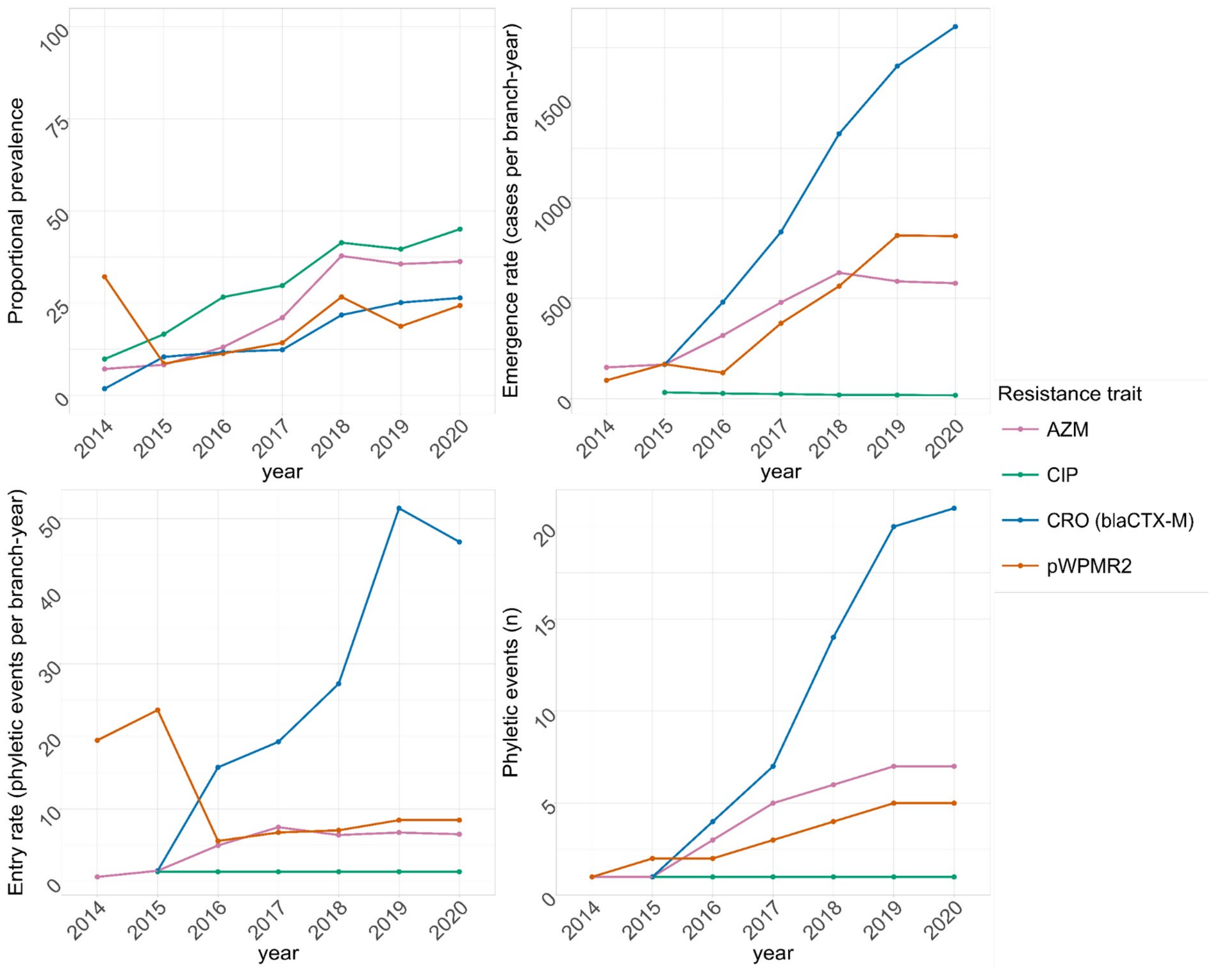


**Supplementary Figure 4.** Time series of the 3,745 *Shigella sonnei* isolates from the UKHSA collection. Starting from the left, the plot shows the trend of i) prevalence ii) Eemergence rate with a filter of minimum five number of descendants from the tMRCA polyphyletic node iii) Eentry rate iv) Number of polyphyletic events detected by ancestral state reconstruction of the four main antimicrobial resistance determinants in *S. sonnei*, plus the presence of a phage plasmid pWPMR2, involved in both survival and increased resistance to third generation cephalosporins. Every dot represents the aggregated values retrieved from each polyphyletic event detected in a specific time frame.

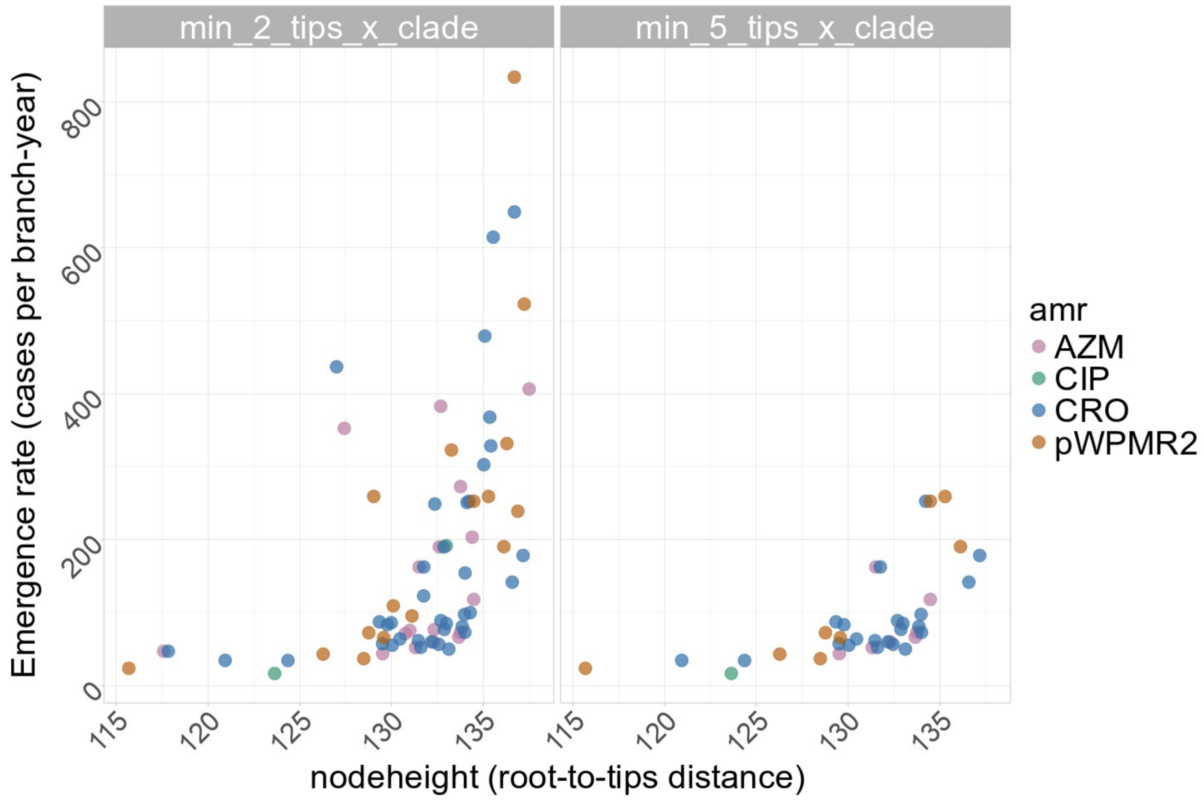


**Supplementary Figure 5.** Plot representing the Emergence rate with the node height of the tMRCA where a polyphyletic event was detected. Each dot represents the Emergence rate of a clade where a polyphyletic event was detected.
